# Paediatric snakebite in Kilifi County, Kenya: A 19-year observational study

**DOI:** 10.1101/2022.11.28.22282866

**Authors:** Michael Abouyannis, Mwanamvua Boga, David Amadi, Nelson Ouma, Amek Nyaguara, Neema Mturi, James A. Berkley, Ifedayo Adetifa, Nicholas R Casewell, David G Lalloo, Mainga Hamaluba

**Affiliations:** KEMRI-Wellcome Trust Research Programme, Centre for Geographic Medicine Research (Coast), Kilifi, Kenya; Centre for Snakebite Research and Interventions. Liverpool School of Tropical Medicine, Liverpool, UK; London School of Hygiene and Tropical Medicine, London, UK; Centre for Tropical Medicine & Global Health, Nuffield Department of Medicine, Oxford, UK

**Author notes:** Corresponding author: (MA). Joint first authors (Michael Abouyannis, Mwanamvua Boga).

## Abstract

**Introduction:** Estimates suggest that one-third of snakebite cases in sub-Saharan Africa affect children. Despite children being at a greater risk of disability and death, there are limited published data describing the burden of paediatric snakebite. This study describes the clinical features, hospital management, population-incidence, and mortality of consecutive paediatric snakebite cases over a 19-year period.

**Methods:** Using the Kilifi Health and Demographic Surveillance System (KHDSS), all children with snakebite presenting to Kilifi County Hospital were identified. All cases were prospectively registered, admitted for at least 24-hours, and managed on a paediatric high dependency unit (HDU). Households within the KHDSS study area have been included in 4-monthly surveillance and verbal autopsy, enabling calculation of population-incidence and mortality. Predictors of severe local tissue damage were identified using a multivariate logistic regression analysis.

**Results:** Between 2003 and 2021, there were 19,798 admissions to the paediatric HDU, of which 619 were due to snakebite. Amongst infants (≤5-years age) the population-incidence of hospital-attended snakebite was 9.8/100,000 person-years; for children aged 6-13-years this was 26.4/100,000 person-years. The number of children attending hospital with snakebite has increased over time. Children aged 8-11 years had the highest incidence. There were six snakebite associated deaths. At admission, low haemoglobin, raised white blood cell count, and an upper limb bite-site were associated with the development of severe local tissue damage.

**Conclusion:** There is a substantial burden of disease due to paediatric snakebite and this has increased in-line with population growth. Most cases present with local rather than systemic envenoming, and many of these receive antivenom. The mortality rate was low, which may reflect the quality of care delivered on the paediatric HDU. There is an urgent need to better define the burden of paediatric snakebite across Africa, to initiate community engagement to prevent snakebites, and to improve treatments for local tissue damage.

## Introduction

Snakebite is a neglected tropical disease that affects 5 million people each year, with the greatest burden falling on rural populations of the tropics and sub-tropics [1]. Snakebite disproportionately affects children, who are at a greater risk of disability and mortality, yet limited published data are available. In sub-Saharan Africa it has been estimated that 30% of people affected by snakebite are children [2]. Typical activities that may bring children into contact with snakes include outdoor play, agricultural work, and walking to school. The burden of snakebite in sub-Saharan Africa has been estimated at over 1 million DALYs per year [3]. Much of this is accounted for by deaths and limb amputations in the young who disproportionately contribute to disability adjusted life years and years of life lost [3].

Younger people are twice as likely to need antivenom treatment following snakebite, and have an increased risk of death [4–6]. Despite this high burden of disease, limited studies describe the burden of paediatric snakebite in sub-Saharan Africa. Two observational studies in South Africa (with samples sizes of 51 and 72 children) identified a substantial burden of painful progressive swelling, with one in four cases undergoing debridement or fasciotomy [7,8]. In a retrospective study of 28 consecutive cases of paediatric snakebite in The Gambia, the mortality rate was 14% [9]. In a community based survey conducted in Kilifi County, Kenya, in 1994, 46% of snakebite cases occurred in children or young adults [10].

Kilifi County is situated on the coast of Kenya and is the site of the KEMRI-Wellcome Trust Research Programme. The Kilifi Health and Demographic Surveillance System (KHDSS) is a longitudinal community- and hospital-based surveillance system [11] that has recorded data, and post-discharge outcomes, on paediatric snakebite victims admitted to hospital since 2003. In this study, comprehensive surveillance and clinical data have been gathered that detail the clinical features, outcomes, and epidemiology of paediatric snakebite in this rural area of Kenya between 2003 and 2021.

## Methods

### Study site

Through a partnership between the Kilifi County Department of Health and the KEMRI-Wellcome Trust Research Programme, clinical data have been prospectively collected for all cases of snakebite attending Kilifi County Hospital since 2003. Informed written consent was sought from the child’s parents or legal guardian. To avoid missing cases with delayed onset of clinical envenoming, local policy stipulates that all cases of paediatric snakebite are admitted for ≥24-hours observation on the paediatric high dependency unit (HDU). This is regardless of disease severity and even applies to cases with no features of envenoming. The paediatric HDU is funded and staffed by the KEMRI-Wellcome Trust Research Programme and a standardised protocol for the management of snakebite is in place. The resources and quality of care available at this paediatric HDU are higher than typical government healthcare facilities in much of Africa, facilitating an approach to snakebite management that aligns with recommended standards in high-income settings.

Hospital surveillance data are linked to the KHDSS community surveillance data. The study area is 891 km^2^ and Kilifi County Hospital is the only hospital with inpatient paediatric services in the study area [11]. The study area, including all dwellings, has been GPS mapped. In 2021, the KHDSS included 92,063 households and 309,228 residents. Most of the study population reside in rural dwellings and the local economy is predominantly centred on subsistence farming [11].

### Identification and eligibility of cases

All cases of snakebite affecting children aged ≤13 years, attending from January 2003 until December 2021, were eligible for inclusion in this study. At admission and on discharge from Kilifi County Hospital, clinical-research staff prospectively assigned a diagnostic code which was recorded on the KHDSS hospital database. Cases with the following diagnostic codes at admission or discharge were searched for: (1) “snake venom”; (2) “snake bite”; or (3) “acute animal bite”. In addition to these diagnostic fields, free-text fields were searched for the following terms: (1) “snake”; or (2) “venom.” All cases identified through this database search were screened by an academic clinician (MA) and non-snakebite cases, for example a dog bite, were excluded. In cases where the diagnosis of snakebite was uncertain, the paper notes were scrutinised by the study team.

### Data extraction

The following data were prospectively recorded on the KHDSS database at the time of hospital admission by research staff: demographics, date and time of admission, date and time of discharge, weight, admission vital signs, admission Blantyre Coma Scale, diagnosis code, mortality, date of death, full blood count, sodium, potassium, urea, and creatinine.

The original paper case notes were sought for every case and data were extracted by a team of research nurses using a standardised case report form. The following data were extracted: demographics; residence; geographic location of bite, circumstances of bite; date and time of bite; anatomical location of bite; use of traditional treatments; clinical features of local and systemic envenoming; vital signs; full blood count; clotting time; renal function; antivenom administration; indication for antivenom; antivenom adverse events; adjunctive treatments; complications of envenoming; and discharge destination. The antivenom product that was administered was not available in the medical records. However, from the hospital pharmacy records it was possible to identify the antivenom product(s) available during each study year.

Deaths were identified by searching the hospital surveillance system, the paper medical records and the KHDSS community surveillance. Verbal autopsy is routinely conducted for all deaths that occur within the KHDSS study area. This is conducted using the 2007 World Health Organization (WHO) VA tools [12], as described previously [13].

To calculate the population-incidence of hospital-attended snakebite, census data from the KHDSS was used. Full details of this surveillance system have previously been published [11]. Community interviewers visit every household in the study area on a 4-monthly basis. A single resident is interviewed, from whom information pertaining to each resident is collected. The identity of all residents is confirmed and any newly born children, in-migrations, deaths, and out-migrations, since the previous enumeration round, are recorded. Person-years of observation are stratified by sex, age and 41 geographic sub-locations.

### Statistical analysis

Clinical data were described using summary statistics including means, medians, and proportions. Population-incidence of hospital-attended snakebite was calculated with 95% confidence intervals. The hospital surveillance system was not consistently capturing all admissions until 2006, thus, to avoid underestimation, incidence estimates have only been calculated for the period of 2006-2021.

A logistic regression analysis was conducted to identify variables associated with severe local tissue damage. Severe local tissue damage was defined as any case developing local skin necrosis or requiring surgical intervention. The following variables were included in a univariate analysis, and those with a significance value of p ≤0.10 were selected for inclusion in the multivariate analysis: age, site of bite (upper limb or lower limb), time-delay from bite to admission, MUAC (mid-upper arm circumference)-for-age z-score (using the *zscorer* package in R), vital signs on admission (pulse rate, respiratory rate, systolic blood pressure, capillary refill time, axillary temperature, oxygen saturations, and Blantyre Coma Scale), admission full blood count (haemoglobin, white cell count, granulocyte count, lymphocyte count, and platelet count), and admission serum biochemistry (sodium, potassium, and estimated glomerular filtration rate (eGFR)). The eGFR was calculated using the Schwartz equation [14]. Multiple imputation, using the *mice* package in R, was undertaken to replace missing values [15]. R version 4.2.2 (R Foundation for Statistical Computing) was used for all analyses.

### Ethical approvals

Written informed consent, with verbal and written patient information translated to Swahili and Giriama, was obtained from the parent or guardian of children enrolling in to the KHDSS and hospital surveillance studies. Ethical approval to undertake this study, including the extraction of data from the hospital records, was granted by the Kenya Medical Research Institute Scientific Ethics Review Unit [KEMRI SERU] (KEMRI/SERU/CGMR-C/174/3930) and Liverpool School of Tropical Medicine Research Ethics Committee [LSTM REC] (19-064) prior to the study starting.

## Results

### Incidence of hospital-attended paediatric snakebite

During the study period there were 78,684 paediatric admissions to Kilifi County Hospital, and 19,798 admissions to the paediatric HDU. The diagnostic code search of the KHDSS hospital database identified 749 potential paediatric snakebite cases. Following manual review of clinical data, 72 were excluded as they were not cases of snakebite (these were predominantly bites by other animals). Further exclusions were as follows: 18 duplicates, 17 were aged over 13 years, 7 attended prior to 2003, and 16 were cases of venom ophthalmia (resulting from venom spitting, rather than snakebite). Therefore, there were 619 children aged 13-years or under that presented with snakebite to Kilifi County Hospital between January 2003 and December 2021. Snakebite thus represented 3.1% of all admissions to the paediatric HDU over the study period. Details of the proportion of admissions to Kilifi County Hospital for snakebite by year are available in Appendix A. The median age was 9 years (IQR 6-11 years) and 46.5% were female. Most cases were resident in the KHDSS study area (N= 424; 68.5%), and clinical records were available for 482 (77.9%) patients. Table 1 provides an overview of the study population.

**Table 1.** Characteristics of paediatric snakebite cases presenting to Kilifi County Hospital.

| **Hospital surveillance data** | **N=619** |
| --- | --- |
| Median age (IQR), years | 9 (6-11) |
| <1 year, n (%) | 2 (0.3) |
| 1 year, n (%) | 8 (1.3) |
| 2-5 years, n (%) | 132 (21.3) |
| 6-9 years, n (%) | 217 (35.1) |
| 10-13 years, n (%) | 260 (42) |
| Sex (%), females | 288 (46.5) |
| Resident in KHDSS study area | 424 (68.5) |
| Abnormal age specific vital signs (%) |  |
| Tachycardia (age adjusted) | 347 (56.3) |
| Tachypnoea (age adjusted) | 382 (62.1) |
| Hypotension (age adjusted) | 26 (5.4) |
| Capillary refill >2 seconds | 9 (1.5) |
| Hypoxia (saturation <95%) | 15 (2.4) |
| Fever >37.2°C | 163 (26.5) |
| Blantyre coma scale (%) |  |
| 5 | 581 (97.2) |
| 4 | 9 (1.5) |
| 3 | 4 (0.7) |
| 2 | 2 (0.3) |
| 1 | 2 (0.3) |
| Severe local tissue damage (%) | 27 (4.4) |
| Local necrosis | 21 (3.4) |
| Required surgery | 20 (3.2) |
| Debridement | 10 (1.6) |
| Fasciotomy | 8 (1.3) |
| Skin graft | 4 (0.6) |
| Amputation | 3 (0.5) |
| Snakebite associated mortality (%) | 6 (1.0) |
| **Clinical record data, n (%)** | **n=482 (77.9)** |
| Circumstances of bite (%) |  |
| Outside home compound | 134 (42.8) |
| Inside home | 71 (22.7) |
| Walking on footpath | 52 (16.6) |
| Farming | 30 (9.6) |
| Walking in bushland | 15 (4.8) |
| At school | 6 (1.9) |
| Hunting for small mammals in burrows | 5 (1.6) |
| Missing | 169 |
| Anatomical site of bite (%) |  |
| Foot | 216 (51.6) |
| Leg | 142 (33.9) |
| Hand | 43 (10.3) |
| Arm | 16 (3.8) |
| Other | 2 (0.5) |
| Missing | 63 |
| Traditional therapies (%) |  |
| Black stone | 114 (23.7) |
| Herbal medicine | 43 (8.9) |
| Puncture to bite site | 50 (10.4) |
| Torniquet | 49 (10.2) |
| Suction | 3 (0.6) |
| Clinical features (%) |  |
| Swelling | 409 (84.9) |
| Blisters | 42 (8.7) |
| Bite site bleeding | 20 (4.1) |
| Bite site infection | 9 (1.9) |
| Systemic bleeding | 7 (1.5) |
| Neurotoxicity | 3 (0.6) |
| No features of envenoming | 51 (11.9) |
| Clinical laboratory (%) * |  |
| Anaemia | 11 (2.7) |
| Leukocytosis | 137 (32.7) |
| Granulocytosis | 43 (57.3) |
| Lymphocytosis | 4 (2.7) |
| Thrombocytosis | 11 (2.7) |
| Thrombocytopaenia | 22 (5.3) |
| Hyponatraemia | 39 (14.0) |
| Hypernatraemia | 9 (3.2) |
| Hypokalaemia | 5 (1.8) |
| Hyperkalaemia | 21 (7.5) |
| eGFR <90 ml/min/1.73 m<sup>2</sup> | 8 (4.8) |
| Antibiotic treatment (%) | 305 (63.3) |
| Cloxacillin | 268 (55.6) |
| Gentamicin | 111 (23) |
| Metronidazole | 58 (12) |
| Ceftriaxone | 20 (4.1) |
| Penicillin | 14 (2.9) |
| Ampicillin | 12 (2.5) |
| Chloramphenicol | 4 (0.8) |
| Other | 9 (1.9) |
| Treatments for acute allergic reactions (%) |  |
| Intravenous chlorphenamine | 20 (16.4) |
| Intravenous hydrocortisone | 19 (15.6) |
| Subcutaneous adrenalin | 7 (5.7) |

eGFR: estimated glomerular filtration rate; IQR: inter-quartile range; KHDSS: Kilifi Health and Demographic Surveillance System.
\*The numbers of missing results by laboratory assay were as follows: haemoglobin, 208 (34.1%); leukocyte count, 199 (32.1%); granulocyte count, 510 (87.9%); lymphocyte count, 458 (75.8%); platelet count, 205 (33.1%); serum sodium, 341 (55.1%); serum potassium, 340 (54.9%); and eGFR, 454 (73.3%).

The population-incidence of hospital-attended snakebite was calculated using the number of admissions per year amongst children that resided in the KHDSS study area (numerator) and annual age-specific census data from the KHDSS (denominator). As young infants had a substantially lower risk of snakebite, population-incidence was stratified between the ages 0-5 years and 6-13 years. For children aged ≤5-years, the average population-incidence between 2006 and 2021 was 9.8/100,000 person-years; for children aged 6-13-years, the average population-incidence was 26.4/100,000 person-years. Figure 1 demonstrates the annual population-incidence for infants and older children, with 95% confidence intervals. Although there is variability between study years the incidence remained broadly consistent until 2020 with a decline in 2021. As there has been a substantial increase in the number of people residing in the KHDSS study area, absolute numbers presenting with snakebite have increased over the study period. In 2006 there were 110,906 person-years of follow-up amongst children ≤13 years of age; by 2021 this had increased to 178,300 person-years.

**Figure 1.**
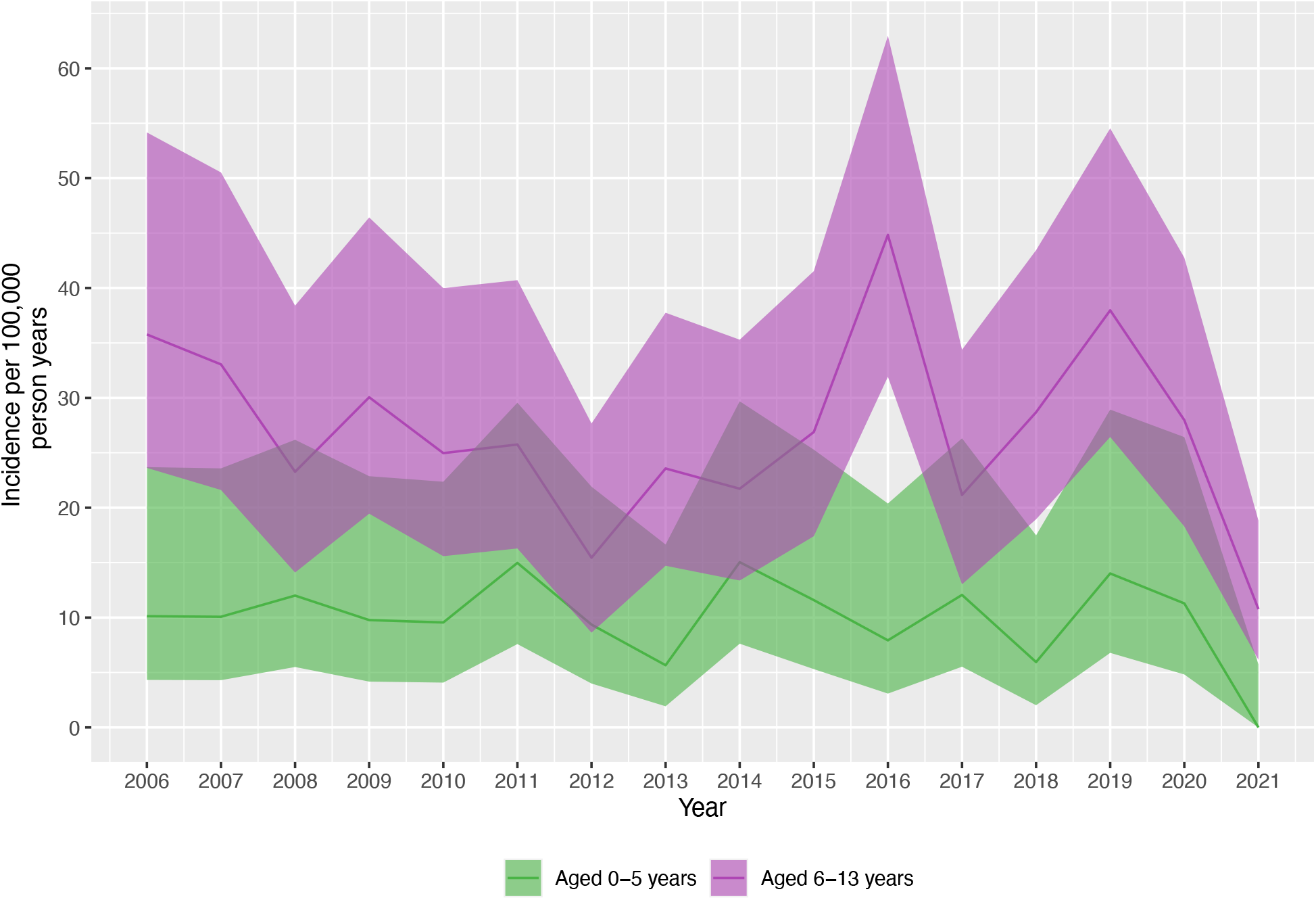
Annual population-incidence of hospital-attended paediatric snakebite.

Population-incidence of hospital-attended snakebite was calculated by year of age, as shown in Figure 2. There was a substantial increase in incidence with age: from 3.6/100,000 person-years at age 1, to 35.5/100,000 person-years at age 11. After age 11, incidence falls, reaching 19.6/100,000 person-years by age 13.

**Figure 2.**
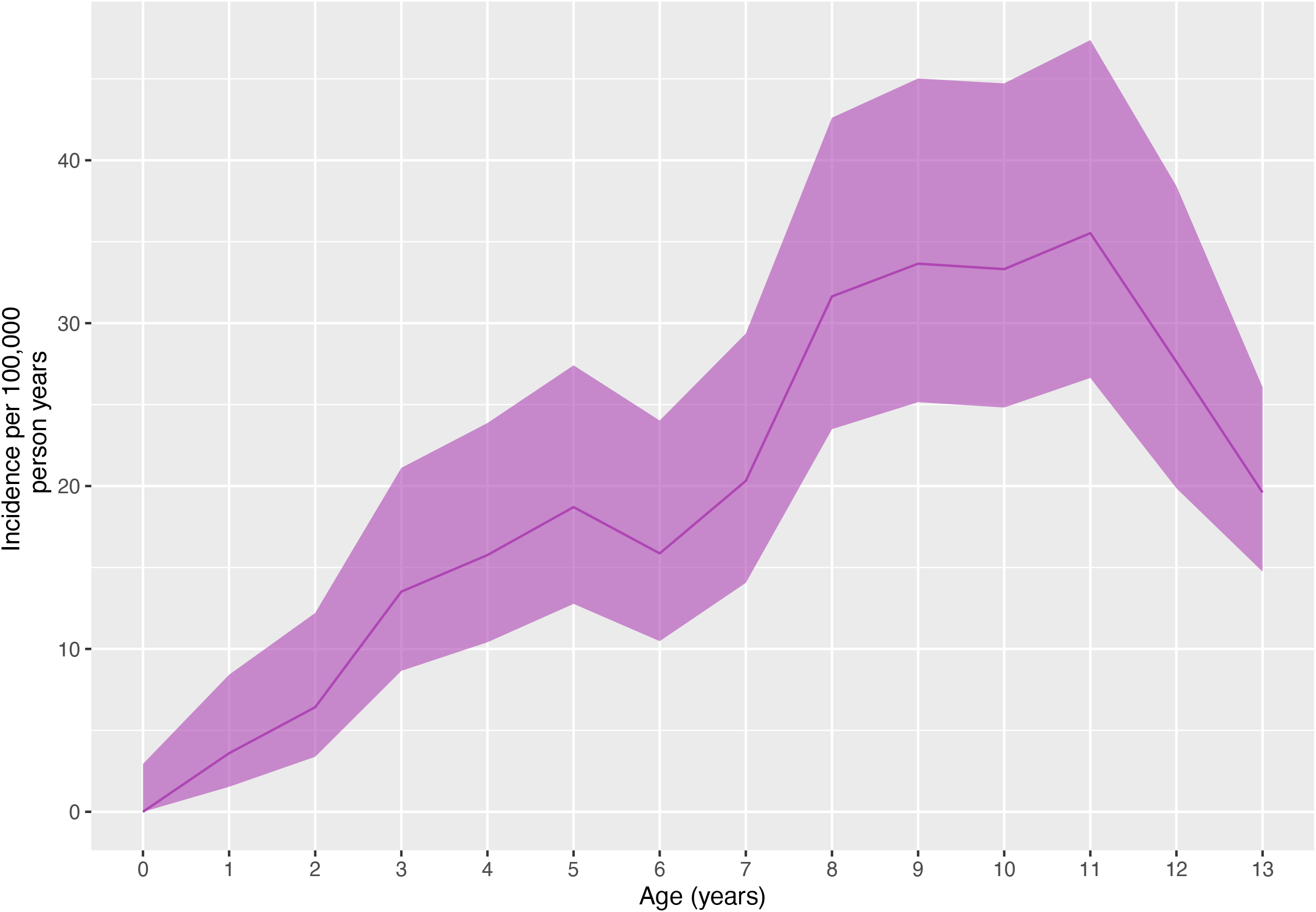
Population incidence of hospital-attended paediatric snakebite by year of age.

### Clinical features

The circumstances of the snakebite were available in the clinical records in 313 (50.6%) cases. The snakebite occurred outdoors and near to the child’s home in 134 (42.8%) cases, 71 (22.7%) occurred in the child’s house, 52 (16.6%) whilst walking on a footpath, 30 (9.6%) whilst farming, 15 (4.8%) in bushland, 6 (1.9%) at school, and 5 (1.6%) whilst hunting for small mammals in burrows. Amongst cases with a documented site of injury (n=419; 67.7%), 216 (51.6%) involved the foot, 142 (33.9%) the leg, 43 (10.3%) the hand, and 16 (3.8%) the arm. Two non-limb bites occurred: to the hip and the external genitalia.

Traditional therapies sought prior to admission included application of a ‘black stone’ in 114 (23.7%) cases, herbal medication in 43 (8.9%) cases, cutting the skin at the bite site in 50 (10.4%) cases, application of a torniquet in 49 (10.2%) of cases, and suction at the bite site in 3 (0.6%) cases. Two or more types of traditional therapy were sought prior to admission in 59 (12.2%) cases (Appendix B). The median time delay from bite until admission was 6.9 hours (IQR 3.3-15.0 hours; range 10 minutes-17 days). The median time delay from admission until administering antivenom was 2.6 hours (IQR 1.2-8.8 hours). Children who had received traditional therapies took an average of two-hours longer to present to hospital, although this difference was not statistically significant (13.1 hours vs 11.3 hours; p=0.39).

Most had local swelling at presentation, being present in 409 (84.9%) cases. Skin blisters were present in 42 (8.7%), bite site bleeding in 20 (4.1%), and bite site infection in 9 (1.9%) cases. There were 29 cases with systemic bleeding, and 3 with neurotoxic envenoming. There were no features of envenoming in 51 cases (11.9%). Age-adjusted tachycardia, hypotension, and tachypnoea were present in 347 (56.3%), 26 (5.4%), and 382 (62.1%) cases, respectively. Fever (>37.2°C) was present in 163 (26.5%) cases, 9 (1.5%) had a prolonged capillary refill time (>2 seconds) and 15 (2.4%) had hypoxia (oxygen saturations <95%). The Blantyre Coma Scale at admission was 5 in 581 (97.2%) cases; 4 in 9 (1.5%) cases; 3 in 4 (0.7%) cases; 2 in 2 (0.3%) cases; and 1 in 2 (0.3%) cases.

Full blood count and serum biochemistry were routinely undertaken for all children presenting with snakebite. Full blood count data demonstrated anaemia (haemoglobin <8.2 g/dL) in 11 (2.7%) cases, leukocytosis in 137 (32.7%), granulocytosis in 43 (57.3%), lymphocytosis in 4 (2.7%), thrombocytosis in 11 (2.7%), and thrombocytopaenia in 22 (5.3%) cases. Blood biochemistry showed hyponatraemia in 39 (14.0%) cases, hypernatraemia in 9 (3.2%), hyperkalaemia in 21 (7.5%), hypokalaemia in 5 (1.8%), and reduced estimated glomerular filtration rate (eGFR) in 8 (4.8%) cases.

### Hospital management

Antivenom was administered to 122 (25.3%) children with snakebite (Table 2). Stratifying by age, antivenom was received by 38 of 115 (33.0%) children aged ≤5-years, and 84 of 367 (22.9%) children aged 6-13-years. A single vial was administered to 74 (67.9%) children, 27 (24.8%) children received two vials, and 8 (7.3%) children received three or more vials. It is estimated that 75 children received Fav-Afrique (Sanofi Pasteur), 12 received ASNA (Bharat Serums and Vaccines), 10 received Snake Venom Antiserum African (VINS Bioproducts), and 25 received Inoserp Pan-Africa (Inosan Biopharma) polyvalent antivenoms (Table 2).

**Table 2.**
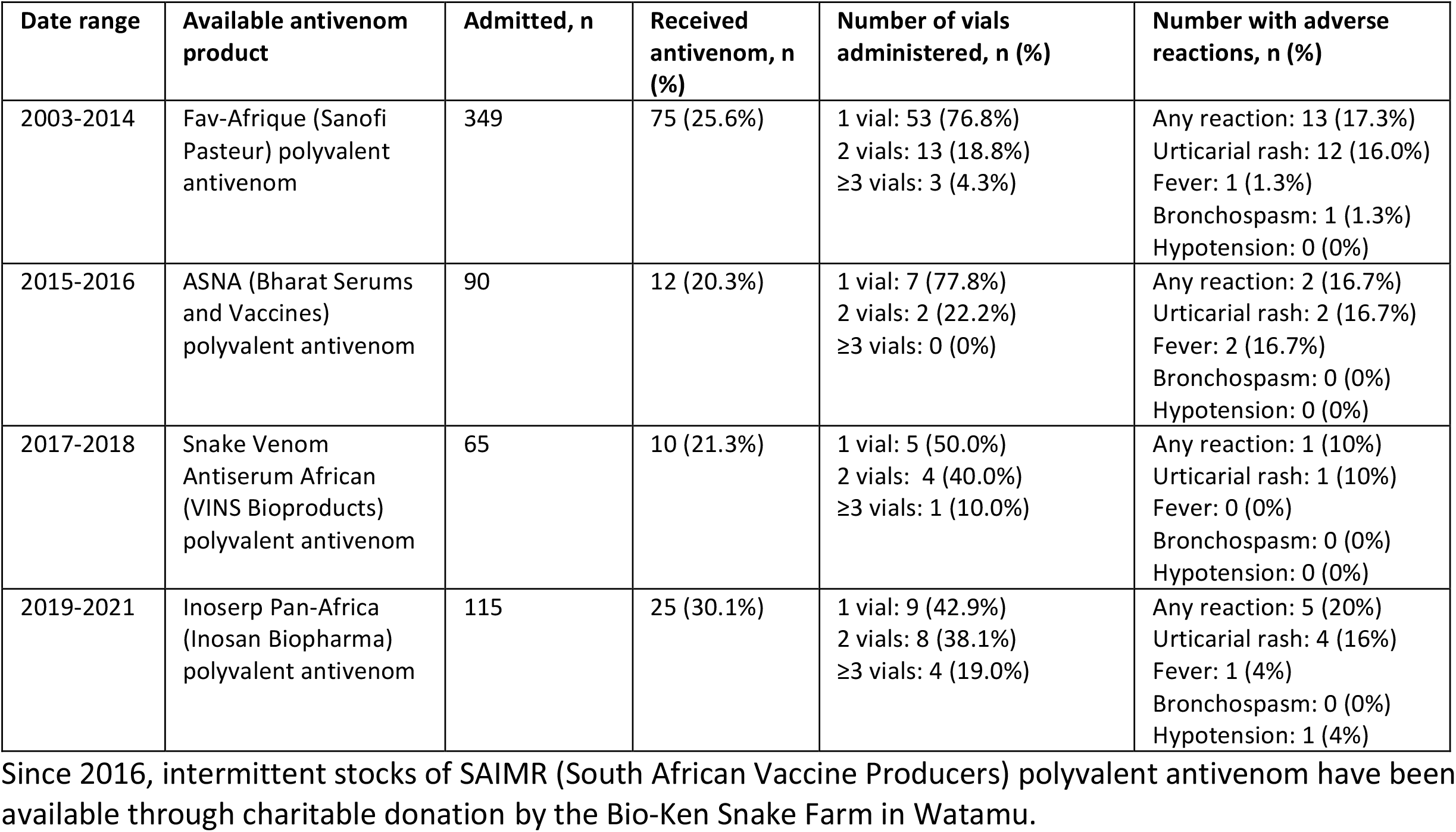
Antivenom products administered, numbers of vials, and rates of acute allergic reactions.

Antivenom was administered for limb swelling in 67 (54.9%) cases, and swelling was the sole indication in 41 (33.6%) cases (Figure 3). Other indications for antivenom included: bleeding in 29 (23.8%) cases, skin blistering in 24 (19.7%), hypotension in 6 (4.9%), restlessness in 13 (10.7%), vomiting in 6 (4.9%), excess salivation in 5 (4.1%), and neurotoxicity in 2 (1.6%) cases (there was a 3^rd^ case of neurotoxicity where it was unknown whether antivenom was administered).

**Figure 3.**
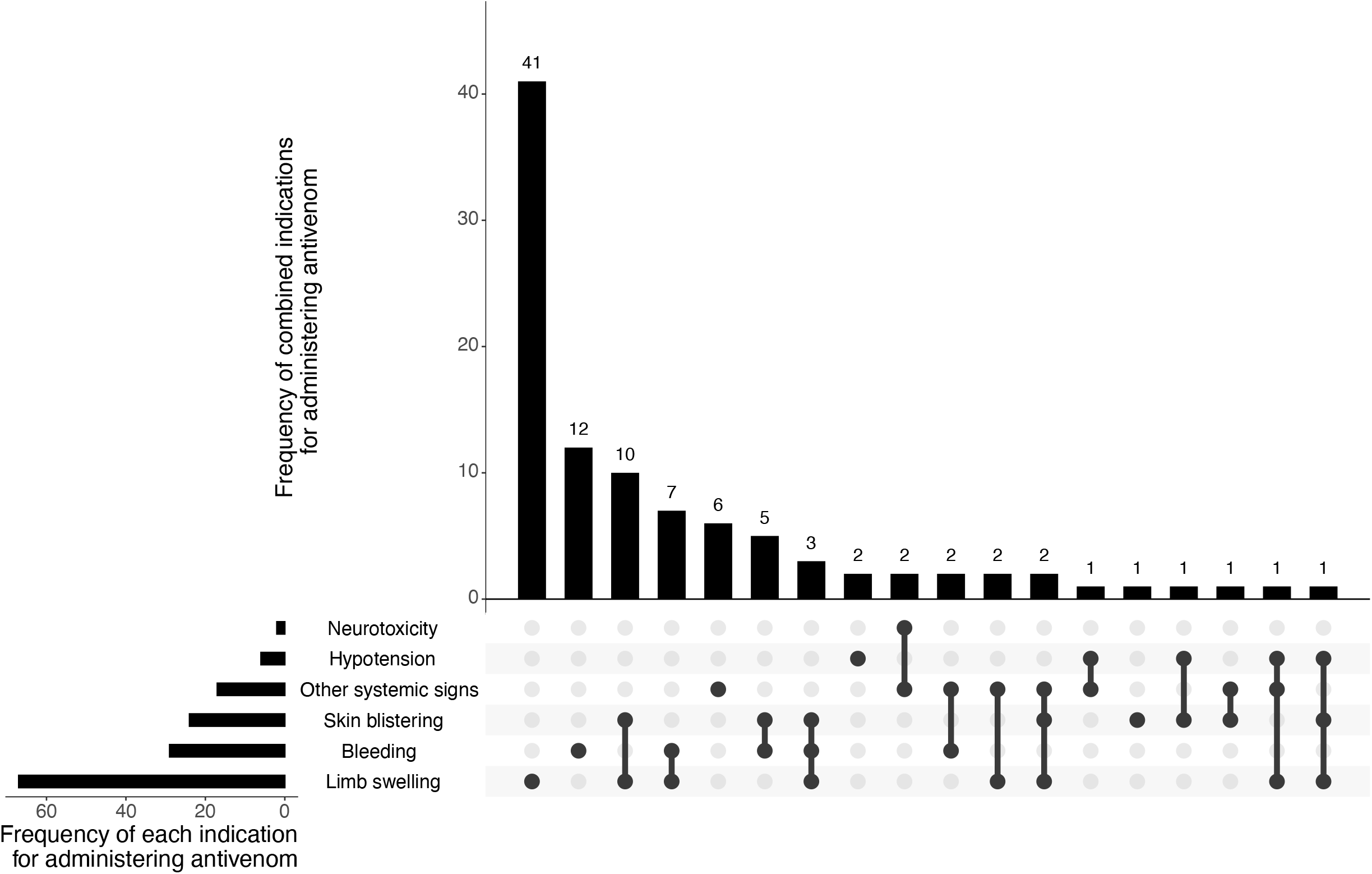
Upset plot of indications for administering antivenom to children with snakebite at Kilifi County Hospital. * Other systemic signs include excess salivation, vomiting, and restlessness

An acute allergic reaction to antivenom occurred in 21 (17.2%) cases (Table 2). A rash occurred in 19 (15.6%) cases, fever in 4 (3.3%), anaphylactic bronchospasm in one (0.8%), and anaphylactic hypotension in one (0.8%) case. The case with anaphylactic hypotension subsequently died due to this complication (Table 3). Amongst children receiving antivenom, 7 (5.7%) received subcutaneous adrenaline, 19 (15.6%) received intravenous hydrocortisone, and 20 (16.4%) received chlorphenamine.

**Table 3.** Characteristics of snakebite associated paediatric deaths.

| Age category (years) | Year of bite | Location of death | Time from bite until attended hospital (hours) | Time from admission to death (days) | Anatomical site of the snakebite | Circumstances of the snakebite | Traditional therapies received | Clinical syndrome of envenoming | Pulse rate on admission (beats/minute) | Respiratory rate on admission (breaths/minute) | Oxygen saturation on admission (%) | Systolic blood pressure on admission (mmHg) | Axillary temperature on admission (°C) | Received antivenom | Developed severe local tissue damage | Clinical details | Cause of death (from hospital records and verbal autopsy) |
| --- | --- | --- | --- | --- | --- | --- | --- | --- | --- | --- | --- | --- | --- | --- | --- | --- | --- |
| 0-5 | 2007 | Kilifi County Hospital | 5 | 0 | Left leg | Inside home | None reported | Bite site swelling, hypotension, bradycardia | 139 | 24 | 97 | NA | 35.3 | No | No | Presented in extremis with hypotensive shock, bradycardia, and respiratory distress. Managed with cardiopulmonary resuscitation, adrenaline, and atropine, but died shortly after admission. | Snakebite |
| 0-5 | 2012 | Kilifi County Hospital | 24 | 1 | Right leg | NA | Puncture to bite site | Bite site swelling | 181 | 58 | 99 | 123 | 37.8 | One vial | No | Sustained a snakebite to the toe. Delayed presentation by 1-day as sought cutting to bite site by a traditional healer. On presentation was irritable and unable to eat. Became progressively lethargic and died following a cardiac arrest the day following admission. | Snakebite |
| 6-13 | 2018 | Kilifi County Hospital | NA | 1 | Right arm | NA | None reported | Bite site swelling, blistering, systemic bleeding, coagulopathy, and hypotension | 104 | 23 | 92 | NA | 36.2 | One vial | No | Bite to arm whilst looking for rats in a burrow. Swelling progressed up to the shoulder and developed systemic bleeding. Underwent fasciotomy for presumed compartment syndrome. Developed uncontrollable haemorrhage and hypotensive shock intra-operatively. Transfused with red blood cells and fresh frozen plasma but developed cardiopulmonary arrest and died. | Haemorrhage secondary to surgical fasciotomy for snakebite |
| 6-13 | 2019 | Kilifi County Hospital | NA | 0 | NA | NA | NA | Neurotoxicity | 66 | 20 | 89 | NA | 39.0 | NA | NA | Bitten by a long brown snake and within hours developed agitation, excessive salivation, difficulty in breathing, and confusion. Initially presented to a local dispensary and then referred to Kilifi County Hospital. Lost consciousness prior to admission and died on the day of admission. | Snakebite |
| 6-13 | 2020 | Kilifi County Hospital | 12 | 1 | Right foot | While walking on a footpath | None reported | Bite site swelling, and neurotoxicity | 122 | 20 | 100 | 112 | 33.9 | Two vials | No | Snakebite to the lower limb was followed by difficulty in swallowing and blurred vision. At hospital developed respiratory arrest which was managed with mechanical ventilation. The following day spontaneous breathing returned, and mechanical ventilation was ceased. Subsequently developed copious haematemesis followed by cardiac arrest. Managed with cardiopulmonary resuscitation and adrenaline but died shortly after. | Snakebite |
| 6-13 | 2020 | Kilifi County Hospital | NA | 0 | Unspecified lower limb | Farming | NA | Bite site swelling | 116 | 24 | 100 | NA | 33.7 | NA | NA | Initially presented to a dispensary following snakebite to the lower limb and was then transferred to Kilifi County Hospital. At admission there was severe swelling up to mid-thigh, bleeding from the bite site, agitation, and confusion. Following two doses of antivenom developed new onset hypotension, tachypnoea, chest pain, and dizziness, which was thought to be anaphylaxis and adrenaline was administered. Condition progressed to cardiopulmonary arrest which did not respond to further doses of adrenaline and manual ventilation. | Antivenom associated anaphylaxis |

Antibiotics were administered to 305 (63.3%) children with snakebite. Cloxacillin was given to 268 (55.6%) children, gentamicin to 111 (23.0%), metronidazole to 59 (12.2%), ceftriaxone to 20 (4.1%), penicillin to 14 (2.9%), ampicillin to 12 (2.5%), and chloramphenicol to 4 (0.8%) children.

The mean duration of hospital stay was 6.3 days (SD 17.6 days). The mean duration of hospital stay was significantly prolonged (51.9 days vs 4.1 days; p=0.0002) in cases with severe local tissue damage (defined as developing skin necrosis or undergoing local surgery). The total number of bed-days for paediatric snakebite admissions were 197.6 days per year.

### Predictors of severe local tissue damage

Overall, 27 cases (4.4%) developed severe local tissue damage. Necrosis at the site of the bite developed in 21 cases (3.4%), and 20 cases (3.2%) required surgery. Ten cases underwent debridement, eight had a fasciotomy, four underwent skin grafting, and three had an amputation. Following multiple imputation, the following covariates were assessed in a univariate logistic regression analysis to identify factors associated with severe local tissue damage: age, MUAC-for-age z-score, site of bite (upper vs lower limb), time from bite to admission, vital signs on admission (axillary temperature, pulse rate, respiratory rate, systolic blood pressure, capillary refill time in seconds, and oxygen saturations), serum sodium, serum potassium, eGFR <90 ml/min/1.73 m^2^, white blood cell count, platelet count, and haemoglobin on admission. In the univariate analysis the following covariates were identified with p≤0.10: upper limb bite (OR 3.56; 95% CI 1.49-8.52; p=0.01); haemoglobin (OR 0.71; 95% CI 0.58-0.87; p<0.01); white blood cell count (OR 1.03; 95% CI 1.01-1.04; p<0.01); pulse rate (OR 1.02; 95% CI 1.00-1.03; p=0.04); and systolic blood pressure (OR 1.03; 95% CI 1.00-1.06; p=0.09) (Table 4). These variables were entered into the multivariate logistic regression analysis in which low haemoglobin (OR 0.68; 95% CI 0.52-0.90; p=0.01), raised white cell count (OR 1.03; 95% CI 1.01-1.05; p=0.01), an upper limb bite site (OR 3.02; 95% CI 1.11-8.19; p=0.03), and increased systolic blood pressure (OR 1.04; 1.00-1.08; p=0.04) were identified as significant predictors (Table 4). Amongst the 59 children with an upper limb bite site, 8 (13.6%) developed severe local tissue damage, compared with 13 (3.6%) of the 357 cases with a lower limb bite site. When the location of bite was categorised to separate bites to the hand from bites to the arm, there remained a statistically significant increased odds of severe local tissue damage for arm (OR 5.37; 95% CI 1.45-19.9; p=0.012) and hand bites (OR 4.51; 95% CI 1.27-16.1; p=0.021), compared to bites sustained to the feet (Appendix C).

**Table 4.** Univariate and multivariate logistic regression analysis of predictors of severe local tissue damage.

| Covariate | All cases | None-severe local tissue damage | Severe local tissue damage | Missing, n (%) | Univariate analysis |  | Multivariate analysis |  |
| --- | --- | --- | --- | --- | --- | --- | --- | --- |
|  |  |  |  |  | OR (95% CI) | p-value | OR (95% CI) | p-value |
| Age, mean (SD) | 8.4 (3.3) | 8.4 (3.3) | 8.5 (3.3) | 0 (0.0) | 1.01 (0.90-1.14) | 0.86 | - | - |
| MUAC-for-age z-score, mean (SD) | -1.3 (1.1) | -1.3 (1.1) | -1.1 (1.1) | 49 (7.9) | 1.13 (0.75-1.68) | 0.56 | - | - |
| Upper limb (%) | 14.2 | 13.2 | 35.8 | 203 (32.8) | 3.56 (1.49-8.52) | 0.01 | 3.02 (1.11-8.19) | 0.03 |
| >12-hour delay bite to admission (%) | 31.7 | 31.8 | 30.2 | 194 (31.3) | 0.91 (0.31-2.67) | 0.86 | - | - |
| Pulse rate, mean (SD) | 115.6 (23.4) | 115.2 (23.1) | 124.5 (27.2) | 3 (0.5) | 1.02 (1.00-1.03) | 0.04 | 1.01 (0.99-1.03) | 0.41 |
| Respiratory rate, mean (SD) | 28.1 (6.7) | 28 (6.7) | 29.4 (5.9) | 4 (0.6) | 1.03 (0.98-1.09) | 0.29 | - | - |
| Systolic blood pressure, mean (SD) | 112.6 (14.9) | 112.4 (14.9) | 118.1 (15.7) | 135 (21.8) | 1.03 (1.00-1.06) | 0.09 | 1.04 (1.00-1.08) | 0.04 |
| Capillary refill time >2 seconds (%) | 1.6 | 1.5 | 0 | 29 (4.7) | 3.99 (0.56-28.65) | 0.17 | - | - |
| Axillary temperature, mean (SD) | 36.8 (0.9) | 36.8 (0.9) | 36.7 (0.9) | 4 (0.6) | 0.96 (0.67-1.36) | 0.80 | - | - |
| Oxygen saturation, mean (SD) | 98.8 (2.5) | 98.8 (2.4) | 98.1 (3.1) | 4 (0.6) | 0.94 (0.86-1.03) | 0.20 | - | - |
| BCS <5 (%) | 2.9 | 2.7 | 0 | 21 (3.4) | 2.85 (0.62-13.20) | 0.18 | - | - |
| Haemoglobin, mean (SD) | 11.6 (1.6) | 11.7 (1.5) | 10.5 (2.7) | 208 (34.1) | 0.71 (0.58-0.87) | 0.00 | 0.68 (0.52-0.90) | 0.01 |
| WCC, mean (SD) | 13.2 (14.4) | 12.5 (11.5) | 27.4 (40) | 199 (32.1) | 1.03 (1.01-1.04) | 0.00 | 1.03 (1.01-1.05) | 0.01 |
| Granulocyte count, mean (SD) | 57.8 (22.8) | 57.5 (22.7) | 64.9 (24.2) | 510 (87.9) | 1.02 (0.96-1.10) | 0.45 | - | - |
| Lymphocyte count, mean (SD) | 28.2 (16.6) | 28.4 (16.4) | 22.5 (18.6) | 458 (75.8) | 0.97 (0.93-1.02) | 0.21 | - | - |
| Platelet count, mean (SD) | 330.3 (115.4) | 330.5 (114.1) | 326.9 (141.7) | 205 (33.1) | 1.00 (1.00-1.00) | 0.94 | - | - |
| Sodium, mean (SD) | 136.9 (4.6) | 136.9 (4.5) | 135.2 (4.4) | 341 (55.1) | 0.93 (0.83-1.03) | 0.15 | - | - |
| Potassium, mean (SD) | 3.6 (0.7) | 3.6 (0.7) | 3.4 (0.6) | 340 (54.9) | 0.69 (0.27-1.78) | 0.42 | - | - |
| AKI (%) | 6.1 | 5.7 | 14.2 | 454 (73.3) | 2.49 (0.67-9.25) | 0.17 | - | - |

AKI: acute kidney injury; BCS: Blantyre coma scale; CI: confidence interval; MUAC: mid-upper arm circumference; OR: odds ratio; SD: standard deviation; WCC: white cell count

### Snakebite associated mortality

Ten of the children in this study have died. Four of these deaths were unrelated to the snakebite and occurred years later during separate hospital episodes. The cause of deaths in these cases were epilepsy, accidental fall, acute respiratory infection associated with HIV/AIDS, and seizures secondary to previous bacterial meningitis, and these deaths occurred 13-, 5-, 5-, and 4-years after the snakebite incident, respectively. Of the six snakebite associated deaths, two were due to neurotoxic envenoming and two were iatrogenic (one due to antivenom anaphylaxis, and one due to complications of fasciotomy surgery). One case presented in extremis with cardiovascular instability and died shortly after admission. In one case of an infant with snakebite there was a general deterioration of uncertain aetiology which culminated in cardio-pulmonary arrest (Table 3). All deaths associated with snakebite occurred within one day of admission.

## Discussion

This 19-year study of consecutive hospital admissions for paediatric snakebite represents one of the most comprehensive analyses of paediatric snakebite in Africa and demonstrates the substantial burden of disease that exists. The population-incidence of hospital-attended paediatric snakebite has been consistently high despite substantial population growth, thus resulting in a growing burden of disease. There was a substantial fall in hospital-attended snakebite incidence in 2021, which is likely to be due to the SARS-CoV-2 pandemic dissuading people from attending hospital [16]. The population-incidence of hospital-attended paediatric snakebite rises with increasing age, and peaks at 8-11 years.

One quarter of paediatric snakebite cases were given antivenom, which was most frequently indicated for local envenoming. Most recently (years 2019-2022), Inoserp Pan-Africa (Inosan Biopharma) polyvalent antivenom has been used, although, in 2022 it was withdrawn from the Kenyan market after failing a risk-benefit assessment conducted by the World Health Organization [17]. Since 2016, intermittent stocks of SAIMR (South African Vaccine Producers) polyvalent antivenom have been available through charitable donation by the Bio-Ken Snake Farm in Watamu, which tends to be reserved for more severe cases, given its evidence of pre-clinical efficacy [18]. Although the frequency of severe allergic reactions was low, there was one case that died as a direct result of antivenom induced anaphylaxis. Despite local envenoming being the most frequent indication for administering antivenom in much of Africa, its effectiveness for this indication is unproven, particularly if it is given late, and clinical trials are urgently needed [19]. Novel oral small molecule therapeutics may hold promise, particularly if they can be administered in rural clinics and thus reduce the time to treatment [20–22].

Bleeding was the most common sign of systemic envenoming. Despite this, measures of coagulopathy, such as the 20-minute whole blood clotting test (20-WBCT), were scarcely documented in the case files. Early detection of coagulopathy is important to avoid delays in administering antivenom for systemic envenoming. Following this study, routine clinical practice at Kilifi County Hospital now includes measuring the 20WBCT in all cases of snakebite.

It was not possible to describe the predominant biting species in this study. It is believed that the puff adder (*Bitis arietans*), spitting cobras (*Naja* spp.), and burrowing asps (*Atractaspis* spp.) are the predominant medically important species in this region, but the relative contribution of these, and other less medically important species, is unknown. Mambas (*Dendroaspis* spp.) and non-spitting cobras (*Naja haje*) are habitual to this region of Kenya. Although there were only three cases of neurotoxic envenoming in this study, two were fatal.

Delayed presentation to hospital was frequent and often prolonged. As most cases resided within the KHDSS study area, which is near to Kilifi County Hospital, it is likely that there is a delay in the decision to attend hospital. Many children had received traditional therapies prior to admission to hospital. The most frequently sought traditional therapy was application of a ‘black stone,’ which is used in many geographic settings despite its lack of efficacy [23].

Most children in this study received antibiotics. The majority had cloxacillin, although broader spectrum agents such as ceftriaxone and gentamicin were also used. Unlike other animal bites, snakebite rarely results in infection and routine antibiotic prophylaxis is not recommended [24].

Severe local tissue damage developed in 4.4% of cases and was often associated with admissions that were weeks or even months long. Low haemoglobin was associated with severe local tissue damage. The direction of causality is uncertain, and it is feasible that children with anaemia have multiple comorbidities, such as malnutrition or chronic malaria, that put them at risk of local tissue damage. Snakebite can cause anaemia as a result of thrombotic microangiopathy, although this is usually associated with thrombocytopaenia, which was uncommon in this study [25]. A raised white cell count on admission was also associated with severe local tissue damage, which has been demonstrated in other settings [26,27]. This is likely to be a bi-directional process, with activation of the innate immune system causing collateral damage at the bite site, and damage of local tissues triggering an immune response. Children that had sustained a snakebite to the upper limb were more likely to develop severe local tissue damage, the reason for which is uncertain. It is regarded that children are at a greater risk of envenoming, compared to adults, as they receive a higher dose of venom relative to their body weight; therefore, it may follow that the small upper limb of a child is particularly at risk. Increasing systolic blood pressure was associated with severe local tissue damage, although the small effect size and borderline level of statistical significance bring into question the clinical significance of this association.

A limitation of this study was that paediatric snakebite cases who did not attend hospital were missed, and therefore the true burden of disease has been under-estimated. A household survey is needed to define the epidemiology of snakebite in Kilifi. A further limitation was the reliance on retrospective data from hospital records. However, documentation on the paediatric HDU tended to be detailed and accurate, with standardised admission and discharge Case Report Forms and contemporaneous daily documentation during admission. All cases were managed on a paediatric HDU which is supported and staffed by the KEMRI-Wellcome Trust Research Programme. Some data points are particularly likely to have been underestimated as they were extracted from the clinical records, such as the estimates on use of traditional therapies.

In conclusion, this study demonstrates the substantial burden of snakebite envenoming amongst children in rural Kenya. This is traumatic for children, interrupts schooling and development, is disruptive for families, and can lead to permanent disability or death. There is an urgent need for improved community awareness, with particular focus on preventative strategies, appropriate first aid, and the importance of early presentation to hospital. Many children in Kilifi receive antivenom for local envenoming, and there is an urgent need to assess whether this is effective.

## Data Availability

The underlying data will be made available as supplementary material

### Appendix A. Proportion of paediatric admissions to Kilifi County Hospital that were due to snakebite

| Year | Paediatric admissions to KCH, n | Paediatric admissions to KCH HDU, n | Paediatric snakebite admissions, n | Percentage of paediatric admissions to KCH due to snakebite | Percentage of paediatric HDU admissions to KCH due to snakebite |
| --- | --- | --- | --- | --- | --- |
| 2003 | 5,590 | 926 | 11* | 0.20% | 1.19% |
| 2004 | 5,066 | 782 | 15* | 0.30% | 1.92% |
| 2005 | 4,749 | 832 | 18* | 0.38% | 2.16% |
| 2006 | 5,016 | 740 | 35 | 0.70% | 4.73% |
| 2007 | 4,498 | 879 | 38 | 0.84% | 4.32% |
| 2008 | 4,401 | 948 | 28 | 0.64% | 2.95% |
| 2009 | 4,866 | 968 | 37 | 0.76% | 3.82% |
| 2010 | 4,395 | 999 | 37 | 0.84% | 3.70% |
| 2011 | 4,255 | 987 | 38 | 0.89% | 3.85% |
| 2012 | 3,661 | 948 | 22 | 0.6% | 2.32% |
| 2013 | 2,851 | 946 | 28 | 0.98% | 2.96% |
| 2014 | 4,095 | 1,178 | 42 | 1.03% | 3.57% |
| 2015 | 4,276 | 1,222 | 44 | 1.03% | 3.60% |
| 2016 | 3,809 | 1,186 | 46 | 1.21% | 3.88% |
| 2017 | 2,291 | 1,141 | 26 | 1.13% | 2.28% |
| 2018 | 3,965 | 1,275 | 39 | 0.98% | 3.06% |
| 2019 | 4,278 | 1,366 | 60 | 1.40% | 4.39% |
| 2020 | 3,460 | 1,348 | 38 | 1.10% | 2.82% |
| 2021 | 3,162 | 1,127 | 17 | 0.54% | 1.51% |
| **Total:** | **78,684** | **19,798** | **619** | - | - |

HDU: high dependency unit; KCH: Kilifi County Hospital.
\* During the period between 2003 and 2005 the numbers of snakebite admissions are underestimated as the hospital surveillance system was not yet fully established.

### Appendix B. Upset plot of traditional therapies sought prior to presenting to hospital amongst children with snakebite at Kilifi County Hospital

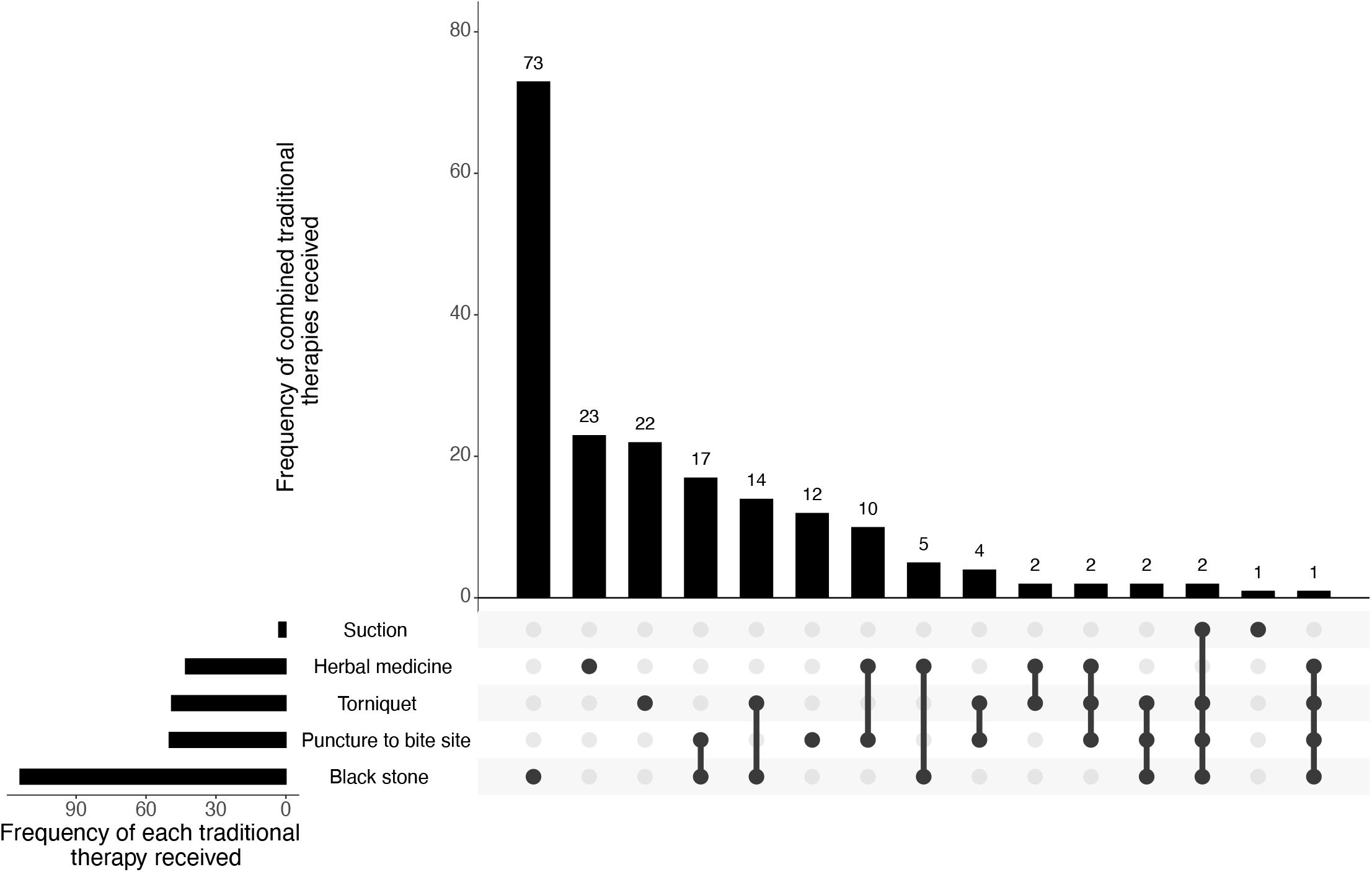

### Appendix C. Multivariate logistic regression analysis of bite site to predict severe local tissue damage amongst children with snakebite at Kilifi County Hospital

| Bite site | OR (95% CI) | p-value |
| --- | --- | --- |
| Arm | 5.37 (1.45-19.91) | 0.012 |
| Hand | 4.51 (1.27-16.09) | 0.021 |
| Leg | 1.40 (0.46-4.26) | 0.557 |

Reference variable: bite to foot.
CI: confidence interval; OR: odds ratio.

